# Genetic Risk and LDL-C Response to Low-Carbohydrate and Low-Fat Diets: A Secondary Analysis of the DIETFITS Trial

**DOI:** 10.64898/2026.11.16.26367963

**Authors:** Alexa Barad, Christopher D. Gardner, Shoa L. Clarke

## Abstract

**Background:** Variability in low-density lipoprotein (LDL-C) response to popular diets is well documented, but the factors contributing to this variability remain unclear. We examined whether genetic predisposition to high LDL-C impacts the effects of healthy low-carbohydrate (HLC) and healthy low-fat (HLF) diets on LDL-C.

**Methods:** The Diet Intervention Examining the Factors Interacting with Treatment Success (DIETFITS) trial randomized 609 adults (ages 18-50 years; body mass index 28-40 kg/m^2^; not on lipid-lowering therapy) to a HLC or HLF diet for 12 months. This secondary analysis included 431 genotyped participants (HLC, n=223; HLF, n=208) with 24-hour recall and LDL-C data at baseline and 6 months. The primary exposure was a genome-wide polygenic score for LDL-C (PGS_LDL_). The outcomes were 6-month change in LDL-C, LDL-C increase versus decrease, and new-onset hypercholesterolemia (LDL-C ≥130 mg/dL). Associations were evaluated using multivariable logistic and linear regression adjusted for baseline LDL-C, weight change, and other factors.

**Results:** Mean baseline LDL-C was 116 mg/dL (SD, 26.1) in the HLC group and 111 mg/dL (SD, 30.6) in the HLF group. On average, LDL-C increased on the HLC diet (+4.7% [SD, 19]) and decreased on the HLF diet (−2.3% [SD, 22]), with substantial variability for both. In the HLC group, higher PGS_LDL_ was associated with risk for LDL-C increase (OR per 1-SD, 1.43; 95% CI, 1.02-2.05) and new hypercholesterolemia (OR per 1-SD, 2.27; 95% CI, 1.18-4.81). Similar effects on LDL-C lowering were not seen for HLF. In both, change in saturated fat intake was associated with change in LDL-C, independent of PGS_LDL_, but this association was modified by PGS_LDL_ only in the HLC arm (*P* for interaction <0.0001).

**Conclusions:** Risk for elevated LDL-C on a HLC diet is greater for individuals with genetic propensity to high LDL-C. This effect may be partly attributed to heightened sensitivity to saturated fat within a low-carbohydrate diet.

**Trial Registration:** clinicaltrials.gov ID: NCT01826591.

## Introduction

The cardiometabolic effects of dietary fat and carbohydrate have long been debated, with common diets centered on modifying these macronutrients (1). Low-carbohydrate diets, which replace carbohydrates predominantly with fat, have gained popularity for weight loss and glycemic control (2, 3). On average, low-carbohydrate diets tend to raise low-density lipoprotein cholesterol (LDL-C), an effect attributed in part to the accompanying increases in saturated fat intake (4). However, this effect varies substantially from person to person, fueling debate over the risks and benefits of this dietary pattern (4). Identifying the factors that drive this heterogeneity is therefore central to understanding the cardiovascular implications of low-carbohydrate diets.

Genetic variation may help explain why individuals differ in their LDL-C response to dietary change. Several dietary trials have evaluated whether individual genetic variants modify this response. Variants in genes involved in lipoprotein metabolism have been associated with differential lipid responses to changes in dietary fat or carbohydrate intake (5–12). However, findings have been inconsistent. For example, some studies have reported greater LDL-C reductions among *APOE* ε4 carriers following low-fat (5–7), or low-saturated fat diets (8), while others have found no such modification (13, 14). Collectively, these studies suggest that genetic variation contributes to lipid responses to diet. However, because LDL-C is a highly polygenic trait, individual variants capture only a small part of the genetic contribution to dietary response, and their modest effects may contribute to the conflicting findings. Whether overall genetic predisposition influences the LDL-C response to dietary interventions remains unclear.

Polygenic scores aggregate the effects of many variants across the genome into a single measure, providing a framework for evaluating whether overall genetic predisposition shapes the lipid response to dietary interventions. We recently showed in the Women’s Health Initiative that the association between saturated fat intake and LDL-C was stronger among individuals with a higher polygenic score for LDL-C (15). In agreement, a smaller observational study suggested that a polygenic score for LDL-C may modify the effects of low-carbohydrate, high-fat diets on LDL-C (16). Importantly, however, this hypothesis has not been tested in a randomized dietary intervention setting. Establishing whether overall genetic predisposition to high LDL-C identifies individuals who experience greater LDL-C increases on a low-carbohydrate diet could help inform the potential cardiovascular implications of this widely adopted dietary pattern.

Here, we use data from the Diet Intervention Examining the Factors Interacting with Treatment Success (DIETFITS) randomized clinical trial (17) and a validated, genome-wide, multi-ancestry polygenic score for LDL-C (PGS_LDL_) (18) to test whether polygenic background influenced the LDL-C response to the healthy low-carbohydrate (HLC) and healthy low-fat (HLF) diet interventions.

## Methods

### Study design

The DIETFITS trial was a single-center, parallel-group randomized controlled trial that enrolled 609 adults aged 18 to 50 years with overweight or obesity (body mass index [BMI], 28-40 kg/m^2^). The trial design, protocol and interventions have been described previously (17, 19). Briefly, participants were randomized to a HLC or HLF diet for 12 months, with change in body weight at 12 months as the primary outcome. Randomization was conducted using a computer-generated allocation sequence (*Blockrand* R package, version 3.4.0). The sequence was generated by a statistician who was not involved in data collection or intervention delivery. Enrollment occurred from January 29, 2013, through April 14, 2015, and final follow-up was completed on May 16, 2016. Key exclusion criteria included uncontrolled hypertension; diabetes or other metabolic disease; cancer; liver, kidney or heart disease; use of lipid-lowering, weight-loss, antihypertensive, antihyperglycemic, or psychiatric medications; current smoker; and pregnancy or lactation. All participants provided written informed consent, and the study was approved by the Stanford University Institutional Review Board.

Participants attended 22 diet-specific behavioral intervention sessions led by registered dietitians. Sessions were held weekly for 8 weeks, biweekly for the subsequent 2 months, every 3 weeks through month 6, and monthly thereafter until the end of the trial. During the initial 8 weeks, participants were instructed to reduce the net carbohydrate or fat intake to 20 grams per day or less, according to their assigned diet, maintaining the lowest level possible for at least one week, followed by gradual increases until each participant found an intake level that they felt they could sustain long term. Both interventions emphasized diet quality, including increased consumption of vegetables and whole foods and reduced intake of processed products, trans fats, added sugars, and refined grains. Neither intervention prescribed explicit energy restriction. Study assessments were conducted at baseline, 3, 6 and 12 months.

The present secondary analysis focused on changes from baseline to 6 months, selected a priori because dietary adherence and participant retention were greater at 6 months than at 12 months (17). The participant flow-chart is shown in **Figure 1**. Of the 609 trial participants, 585 had genotyping data. We excluded related individuals at the second degree or closer (n = 11), those whose genetically inferred ancestry groups were too small to support reliable ancestry-specific PGS_LDL_ normalization (n = 4). We further excluded those with missing baseline or 6-month LDL-C measurements (n = 126) or dietary data (n = 13). The final analytic sample comprised 431 individuals, including 223 randomized to the HLC intervention and 208 randomized to the HLF intervention.

**Figure 1.**
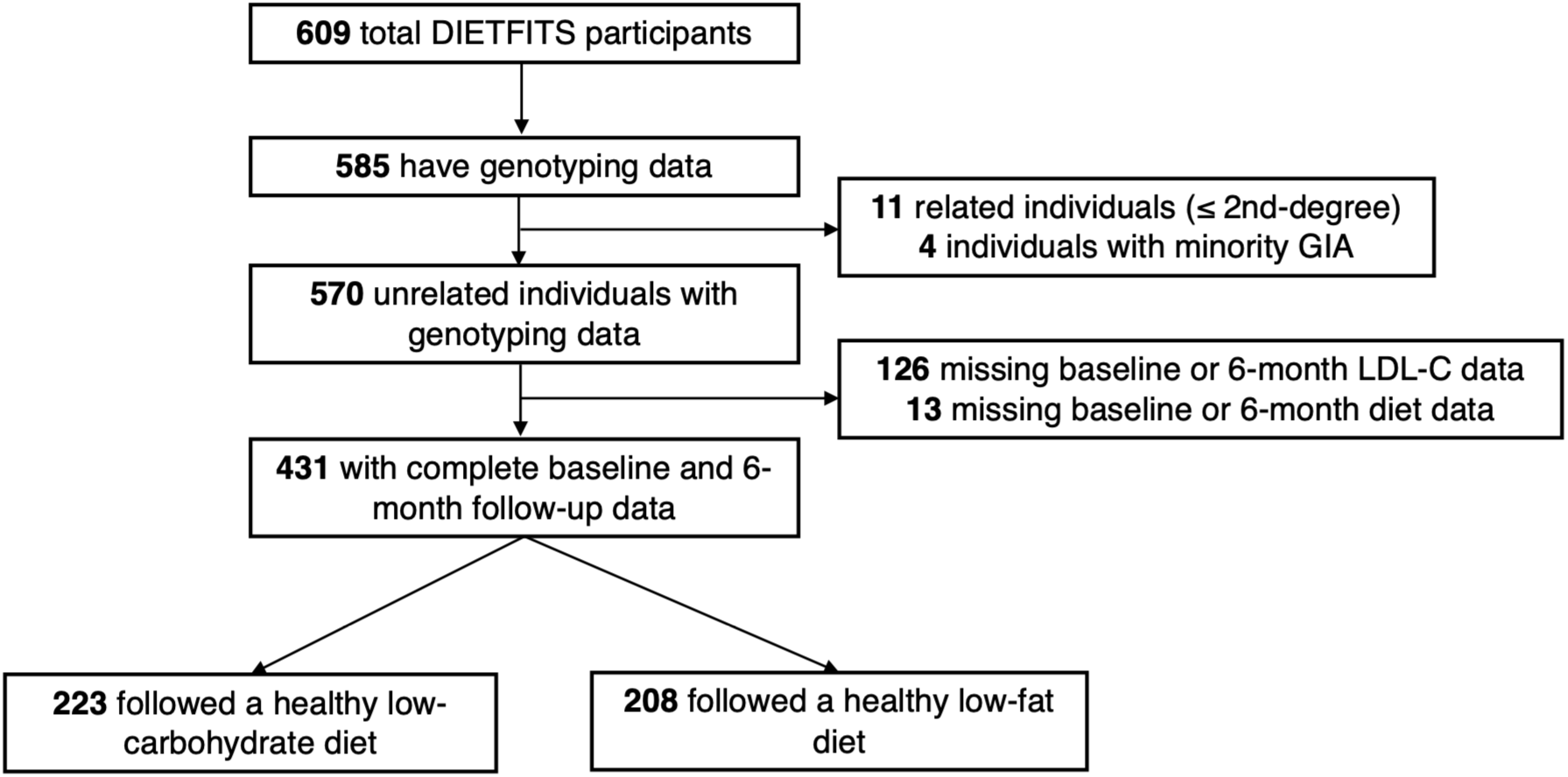
Flowchart of study participants. DIETFITS, Diet Intervention Examining the Factors Interacting with Treatment Success randomized clinical trial; GIA, genetically inferred ancestry; LDL-C, low-density lipoprotein cholesterol.

### Dietary intake and clinical measures

Dietary intake was assessed using three unannounced 24-hour recalls, including two weekdays and one weekend day, using a standardized multiple-pass method (20). Recalls were processed using the Nutrient Data System for Research software (Nutrition Coordinating Center, University of Minnesota, Minneapolis, MN) to obtain nutrient-level data (21). Diet quality was assessed using the Healthy Eating Index-2010 (HEI-2010), calculated for each recall following standard scoring procedures (22, 23). All dietary variables, including macronutrient intake and diet quality scores, were averaged across the three recalls at each time point.

Clinical assessments, including body weight measures and blood collection, were performed by a nurse or phlebotomist at the Stanford Clinical and Translational Research Unit (Stanford University School of Medicine, California) at each study visit. Blood samples were collected after a 10-12 hour fast at each time point. Lipid measurements were performed by the Krauss Laboratory at the Children’s Hospital Oakland Research Institute. Plasma triglycerides, total cholesterol and high-density lipoprotein cholesterol were measured by enzymatic point analysis on a clinical chemistry analyzer (Liasys 330). LDL-C concentrations were calculated using the Friedewald equation (24). Data collectors were blinded to the participants’ assigned diet intervention.

### Genotyping data and polygenic score calculation

Genotyping was performed using the Affymetrix UK Biobank Axiom microarray. Genetic relatedness was estimated using KING (25), and individuals with a kinship coefficient > 0.0884, indicative of a second-degree or closer relatedness, were excluded (**Figure 1**). Imputation was performed using the Trans-Omics for Precision Medicine (TOPMed) imputation server, with TOPMed version r3 as the reference panel (26). Genetic ancestry was inferred using the *pgsc_calc* pipeline (27), which projects principal components and assigns individuals to ancestry groups based on genetic similarity to the 1000 Genomes Project Phase 3 superpopulations (28). We used a previously validated multi-ancestry genome-wide PGS_LDL_ (PGS000889) (18), calculated using *pgsc_calc* (27). Of the 9,009 autosomal variants included in the original score, 8,488 (94.2%) were available in the study cohort and used to calculate the score. The PGS_LDL_ was adjusted for genetic ancestry using the 1000 Genomes reference panel and the eMERGE consortium approach (29), which accounts for differences in score means and variances across ancestry groups. Coefficients are expressed per 1-SD change in the ancestry-normalized PGS_LDL_.

### Statistical analyses

All analyses were stratified by intervention arm. Continuous variables are presented as means and standard deviations, and categorical variables as percentages. Changes in LDL-C and dietary variables from baseline to 6 months were calculated as 6-month minus baseline values. Percent change in LDL-C was calculated as the difference between 6-month and baseline values divided by the baseline value and multiplied by 100. PGS_LDL_ performance in the study population was evaluated using regression coefficients and incremental R² (coefficient of determination), with 95% confidence intervals estimated via percentile bootstrapping. Adequate ancestry normalization of the PGS_LDL_ was assessed using a chi-squared test to confirm that the PGS_LDL_ was not associated with genetically inferred ancestry.

Multivariable logistic regression was used to evaluate the associations between the PGS_LDL_ and 6-month change in LDL-C, categorized as an increase versus decrease in the HLC arm and a decrease versus increase in the HLF arm. Among those with normal LDL-C at baseline, we also evaluated the association between PGS_LDL_ and new-onset hypercholesterolemia, defined as LDL-C ≥130 mg/dL at 6 months. Models were adjusted for age, sex, baseline LDL-C, 6-month change in weight, 6-month change in saturated fat intake, and genetic principal components 1-3.

Multivariable linear regression was used to evaluate the associations of change in saturated fat intake (as percent of total daily energy) and PGS_LDL_ with change in LDL-C (modeled continuously) and to assess whether the PGS_LDL_ modified the association between change in saturated fat intake and change in LDL-C. Additive and interaction models were examined with PGS_LDL_ modeled both continuously and categorically using tertiles. The additive models included main effects for change in saturated fat intake and PGS_LDL_, whereas the interaction models additionally included a multiplicative interaction term between these predictors. Interaction *P-* values were derived from likelihood ratio tests comparing models with and without the interaction term. To characterize the association between change in saturated fat intake and change in LDL-C within each PGS_LDL_ tertile, tertile-specific slopes and 95% confidence intervals were estimated using the *emtrends()* function in the *emmeans* R package (version 2.0.3). All models were adjusted for age, sex, baseline LDL-C, change in weight, and genetic principal components 1-3.

The incremental R^2^ was estimated to quantify the variance in LDL-C response explained by change in saturated fat intake, PGS_LDL_, and their interaction. For each main predictor (change in saturated fat intake and PGS_LDL_), the incremental R^2^ was calculated as the difference in R^2^ between models with and without the predictor of interest, with both models adjusted for the other predictor and base covariates (age, sex, baseline LDL-C, change in weight, and genetic principal components 1-3). For the interaction term, the incremental R^2^ was calculated as the difference in R^2^ between the interaction model and the corresponding additive model. The 95% confidence intervals for incremental R^2^ were estimated via percentile bootstrapping.

We conducted several sensitivity analyses to assess the robustness of our findings. First, we re-ran all models with additional adjustment for changes in diet quality and dietary adherence. Change in diet quality was assessed using a modified HEI-2010 that excluded the fatty acid and solid fat components, which capture saturated fat intake that was already included in the models. Dietary adherence was represented using a binary variable, with high adherence defined as a change in grams of carbohydrate intake (HLC) or fat intake (HLF) below the group-specific median (i.e., a greater decrease) and low adherence defined as a change greater than or equal to the group-specific median. The median 6-month changes, which served as respective cutoffs, were-139.4 grams of carbohydrate in the HLC group and −36.3 grams of fat in the HLF group. Second, we re-ran all models restricted to individuals with genetically inferred European ancestry, as this was the largest ancestry group in which the PGS_LDL_ is known to perform best (18), to assess whether ancestry differences may have influenced the associations. Third, we examined interactions between the PGS_LDL_ and changes in individual saturated fatty acids, stratified by chain length, focusing on saturated fatty acids that contributed more than 1% of total daily energy at baseline. To account for multiple testing burden, a significant interaction for this analysis was defined as a *P* < 0.017 (0.05/3 fatty acids tested). Fourth, we evaluated interactions between the PGS_LDL_ and changes in other macronutrients, including changes in unsaturated fat, carbohydrate and protein. Similarly, a Bonferroni-correction was applied, with significant interactions defined as *P* < 0.0125 (0.05/4 macronutrients). Statistical analyses were performed using R version 4.3.3 (The R Foundation for Statistical Computing). Statistical significance was defined as *P* < 0.05 unless otherwise specified.

## Results

### Cohort characteristics

Of 609 randomized participants, 431 were included in this secondary analysis, with 223 in the HLC group and 208 in the HLF group. Baseline characteristics were broadly similar between diet groups (**Table 1**). The mean age was 40.2 (SD, 6.7) years in the HLC group and 39.2 (SD, 6.7) years in the HLF group, and mean BMI was 33.1 (SD, 3.4) and 33.5 (SD, 3.4) kg/m^2^, respectively. In both groups, more than half of the participants were female, self-identified as White, and had genetically inferred European ancestry, and more than three-quarters had a college degree or higher.

**Table 1.** Cohort characteristics by diet group.^1^.

|  | **Healthy Low-Carbohydrate Diet**<br>**(n = 223)** | **Healthy Low-Fat Diet**<br>**(n = 208)** |
|---|---|---|
| **Sex, n (%)** |  |  |
| Female | 131 (58.7) | 119 (57.2) |
| Male | 92 (41.3) | 89 (42.8) |
| **Age, years** | 40.2 (6.7) | 39.2 (6.7) |
| **Genetically inferred ancestry, n (%)** |  |  |
| European | 145 (65.0) | 127 (61.1) |
| African | 6 (2.7) | 8 (3.8) |
| American | 39 (17.5) | 48 (23.1) |
| East Asian | 19 (8.5) | 12 (5.8) |
| Central/South Asian | 14 (6.3) | 13 (6.2) |
| **Self-reported race/ethnicity, n (%)** |  |  |
| White | 137 (61.4) | 122 (58.7) |
| Black | 7 (3.1) | 6 (2.9) |
| Hispanic | 41 (18.4) | 46 (22.1) |
| Asian | 25 (11.2) | 18 (8.7) |
| Native Hawaiian/Pacific Islander | 2 (0.9) | 1 (0.5) |
| American Indian/Alaskan | 0 (0.0) | 1 (0.5) |
| Other | 11 (4.9) | 14 (6.7) |
| **Highest level of education, n (%)** |  |  |
| < College degree | 48 (21.6) | 47 (22.6) |
| ≥ College degree | 174 (78.4) | 161 (77.4) |
| **Weight, kg** |  |  |
| Baseline | 95.3 (15.5) | 96.8 (14.3) |
| 6 months | 87.4 (14.7) | 90.2 (13.9) |
| Change | -7.9 (6.1) | -6.6 (5.8) |
| **Body mass index, kg/m<sup>2</sup>** |  |  |
| Baseline | 33.1 (3.4) | 33.5 (3.4) |
| 6 months | 30.3 (3.6) | 31.3 (3.6) |
| Change | -2.7 (2.0) | -2.3 (1.9) |
| **LDL-C, mg/dL** |  |  |
| Baseline | 115.7 (26.1) | 111.4 (30.6) |
| 6 months | 119.5 (28.6) | 106.4 (20.3) |
| Change | 3.8 (20.0) | -5.0 (20.2) |

Abbreviations: LDL-C, low density lipoprotein cholesterol.
<sup>1</sup>Values reported as the mean (SD) for continuous variables, or n (percent) for categorical variables.

Detailed dietary intake at baseline and 6 months, as well as changes over this period, are shown in **Supplementary Table 1**. Baseline dietary intake was similar between diet groups, with approximately 44%, 35% and 17% of total daily calories from carbohydrate, fat and protein, respectively, in both. Consistent with diet assignment, at 6 months, mean carbohydrate intake decreased to 26% of calories in the HLC group, and mean total fat intake decreased to 27% of calories in the HLF group.

Mean baseline LDL-C was 116 mg/dL (SD, 26.1) in the HLC group and 111 mg/dL (SD, 30.6) in the HLF group (**Table 1**). The PGS_LDL_ explained 15-17% of the variance in baseline LDL-C (**Supplementary Table 2**), consistent with its expected predictive performance (18). The score was not predictive of genetically inferred ancestry group, supporting adequate ancestry normalization (**Supplementary Table 3**).

### Association between the polygenic score and LDL-C response

At 6 months, LDL-C increased by 4.7% (SD, 19) in the HLC group and decreased by 2.3% (SD, 22) in the HLF group. However, individual LDL-C responses were highly heterogeneous within each diet group. Percent changes in LDL-C ranged from a 26% decrease to a 126% increase in the HLC group and from a 53% decrease to a 125% increase in the HLF group (**Figure 2**).

**Figure 2.**
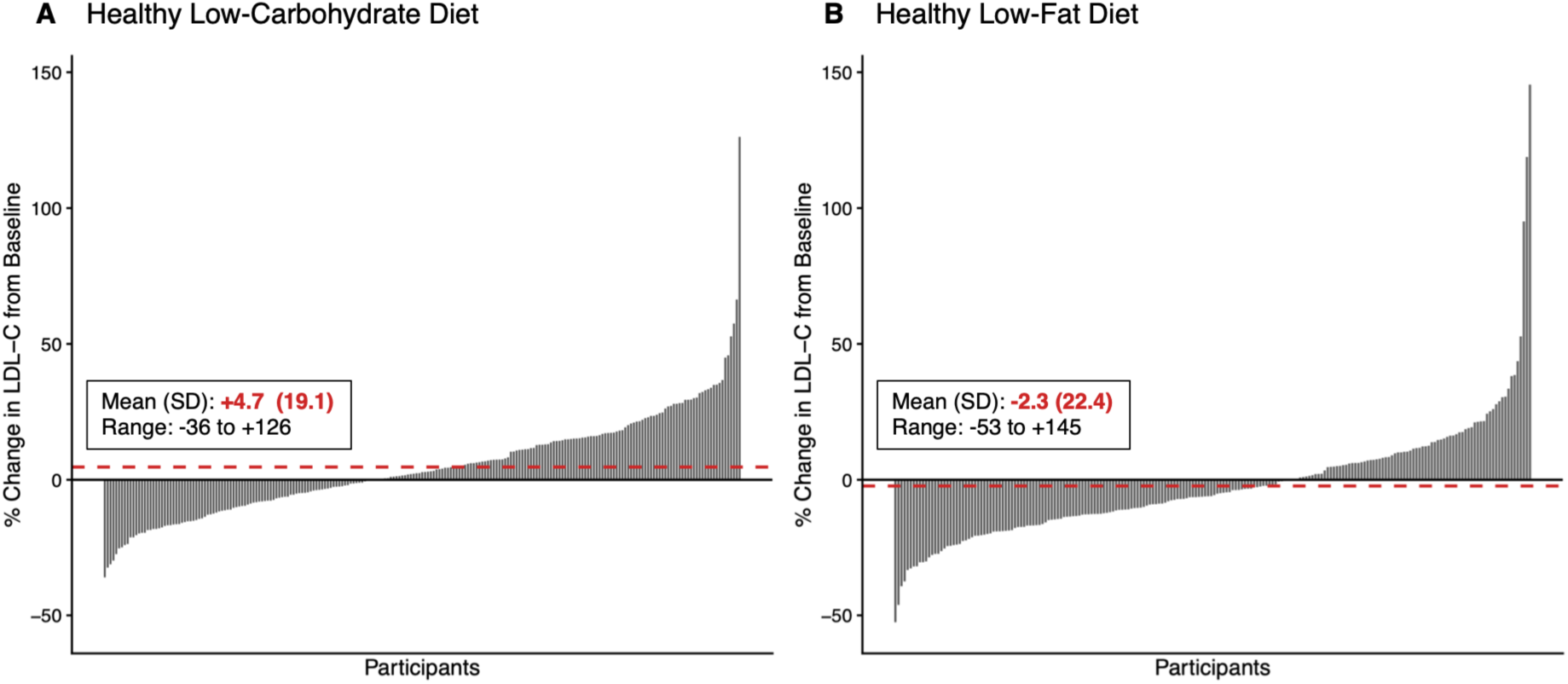
Percent change in LDL-C from baseline to 6 months by diet group. Waterfall plots showing individual-level percent change in low-density lipoprotein cholesterol (LDL-C) from baseline to 6 months among participants randomized to the healthy low-carbohydrate diet (n, 223) (A) and healthy low-fat diet (n, 208) (B). Each bar represents one participant. Red dashed lines represent the mean percent change in LDL-C within each diet group.

We evaluated whether the PGS_LDL_ predicted the direction of LDL-C change in response to each diet, and among participants with normal baseline LDL-C (<130 mg/dL), we assessed whether the PGS_LDL_ predicted the development of hypercholesterolemia. On the HLC diet, 127 (57%) participants experienced an LDL-C increase. The risk for LDL-C increase was 43% higher for each 1-SD increase in PGS_LDL_ (OR, 1.43; 95% CI, 1.02-2.05; *P* = 0.04). Among 151 participants in the HLC group with normal baseline LDL-C (<130 mg/dL), 23 (15.2%) developed hypercholesterolemia. The risk for hypercholesterolemia on the HLC diet increased 2.27-fold for each 1-SD increase in PGS_LDL_ (OR, 2.27; 95% CI, 1.18-4.81; *P* = 0.02). These findings remained consistent after additional adjustment for change in diet quality and dietary adherence, and when analyses were restricted to participants of European ancestry, including models with additional adjustment for these variables in the European ancestry subset (**Supplementary Table 4**).

On the HLF diet, 130 (62.5%) participants experienced an LDL-C decrease, but PGS_LDL_ was not associated with odds of LDL-C decrease (OR, 0.85; 95% CI, 0.59-1.21; *P* = 0.38). Similar nonsignificant findings were observed after further adjustments for change in diet quality and dietary adherence, as well as when analyses were restricted to the European ancestry subset (**Supplementary Table 5**). As expected, new hypercholesterolemia was uncommon on the HLF diet, affecting 8 (5.1%) participants of 157 with normal baseline LDL-C, and the PGS_LDL_ was not associated with this change (OR, 1.4; 95% CI, 0.5-4.9; *P* = 0.52). Of note, given the low incidence of new-onset hypercholesterolemia, this analysis was conducted for comparison purposes only, as the small number of cases limited the ability to meaningfully evaluate this association.

### Interaction between the polygenic score and change in saturated fat intake

Given the well-established effect of saturated fat intake on LDL-C (30, 31), we examined the association between change in saturated fat intake and LDL-C response, and whether PGS_LDL_ modified this association. On average, saturated fat intake increased by 3.2 percentage points of total calories in the HLC group and decreased by 3.6 percentage points in the HLF group (**Supplementary Table 1**). In both groups, change in saturated fat intake was associated with change in LDL-C, independent of PGS_LDL_ (**Table 2**). However, an interaction between change in saturated fat intake and PGS_LDL_ was observed only in the HLC arm. The interaction term explained 3.3% of the variance in LDL-C response, and the PGS_LDL_ main effect and interaction together explained 10.4% of the variance in LDL-C response to the HLC diet (**Table 2**).

**Table 2.** Additive and interaction effects of polygenic score and change in saturated fat intake on 6-month change in LDL-C.^1^.

|  |  | **Healthy Low-Carbohydrate Diet<br>(n = 223)** |  |  | **Healthy Low-Fat Diet<br>(n = 208)** |  |  |
|---|---|---|---|---|---|---|---|
| **Model** | **Independent variable** | **β (SE)** | ***P* value** | **Incremental R<sup>2</sup> (95%CI), %** | **β (SE)** | ***P* value** | **Incremental R<sup>2</sup> (95%CI), %** |
| Additive model | Δ SFA (1 % kcal) | 0.91 (0.28) | 0.002 | 3.8 (0.2, 9.4) | 1.12 (0.33) | 0.0008 | 4.2 (0.3, 11.1) |
|  | PGS<sub>LDL</sub> (1-SD) | 6.04 (1.38) | <0.0001 | 7.1 (1.8, 14.0) | 3.66 (1.35) | 0.007 | 2.7 (0.2, 6.6) |
| Interaction model | Δ SFA (1 % kcal) | 1.03 (0.28) | 0.0003 | - | 1.23 (0.34) | 0.0003 | - |
|  | PGS<sub>LDL</sub> (1-SD) | 3.17 (1.65) | 0.056 | - | 5.93 (1.98) | 0.003 | - |
|  | PGS<sub>LDL</sub> x Δ SFA | 0.69 (0.23) | 0.003 | 3.3 (0.5, 9.3) | 0.56 (0.36) | 0.12 | 0.9 (0, 4.3) |

Abbreviations: LDL-C, low density lipoprotein cholesterol; PGS<sub>LDL</sub> , polygenic score for LDL-C, Δ SFA, change in saturated fat intake (as  percent of total daily energy) from baseline to 6 months.
<sup>1</sup> The β coefficients (SE) represent the change in LDL-C associated with a 1-SD increase in the PGS<sub>LDL</sub> or a 1-percentage-point increase in SFA from baseline to 6 months. All models were adjusted for age, sex, 6-month weight change, baseline LDL-C, and genetic principal components 1-3 (base covariates). Incremental R<sup>2</sup> values represent the additional percent of variance explained beyond the base covariate model. In additive models, incremental R<sup>2</sup> reflects the increase in variance explained by adding each predictor to a model already containing the other predictor and the base covariates. In interaction models, incremental R<sup>2</sup> reflects the additional variance explained by inclusion of the interaction term. The 95% CIs were estimated using bootstrapping.

The observed interaction between change in saturated fat intake and PGS_LDL_ in the HLC group was robust across several sensitivity analyses. First, the interaction remained significant after additional adjustment for change in diet quality and dietary adherence (Interaction β [SE] = 0.68 [0.23]; *P* = 0.003; **Supplementary Table 6**). Second, to account for potential ancestry-related differences, we additionally adjusted for ancestry-by-PGS_LDL_ and ancestry-by-change in saturated fat intake interaction terms, which had minimal impact on the change in saturated fat intake interaction-by-PGS_LDL_ estimate (Interaction β [SE] = 0.59 [0.24]; *P* = 0.01). Findings were also consistent when analyses were restricted to participants of European ancestry (**Supplementary Table 7**). The same sensitivity analyses in the HLF group consistently provided no evidence of an interaction (**Supplementary Tables 6-7**).

We then evaluated the change in saturated fat-by-PGS_LDL_ interaction using PGS_LDL_ tertiles. Consistent with the continuous analysis, the interaction between PGS_LDL_ tertiles and change in saturated fat intake was significant in the HLC group only (HLC *P* interaction < 0.0001 and HLF *P* interaction = 0.47; **Figure 3**). In the HLC group, the effect of increased saturated fat intake on LDL-C increase was attenuated in the lowest PGS_LDL_ tertile and amplified in the highest PGS_LDL_ tertile (**Figure 3A**). In the HLF group, the effects of decreased saturated fat intake on LDL-C did not differ across PGS_LDL_ tertiles (**Figure 3B**). These results were consistent in sex-stratified analyses (**Figure 3**).

**Figure 3.**
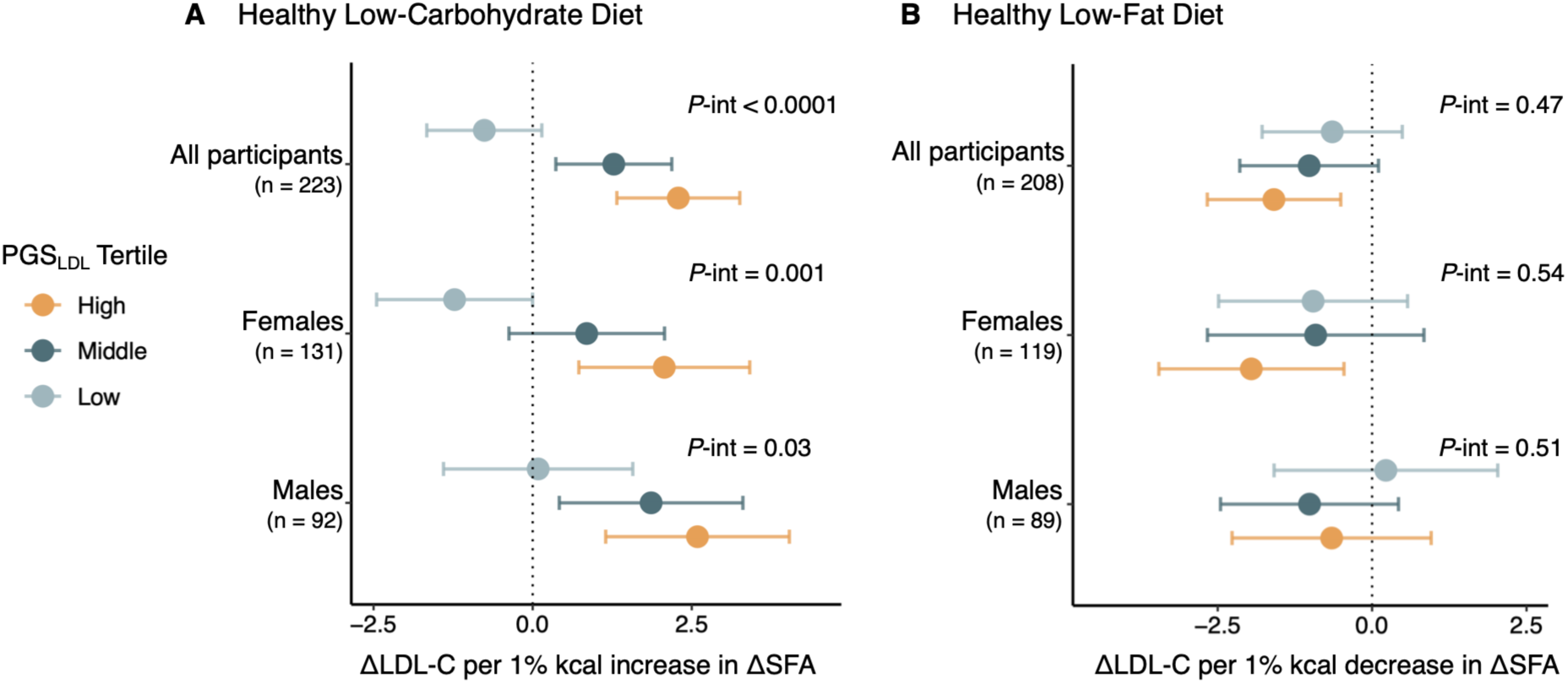
Association of change in saturated fat intake with change in LDL-C according to polygenic score tertile by diet group. Estimated change in low-density lipoprotein cholesterol (LDL-C) per change in 1% of total energy intake from saturated fat (SFA) according to polygenic score (PGS_LDL_) tertile among participants randomized to the healthy low-carbohydrate diet (A) and healthy low-fat diet (B). Analyses are shown for all participants and stratified by sex. Points represent β coefficients and error bars denote 95% CIs from multivariable linear regression models adjusted for age, baseline LDL-C, 6-month weight change, and genetic principal components 1-3. *P* values (*P*-int) indicate interaction between change in saturated fat intake and PGS_LDL_ tertile.

Lastly, in the HLC group, we further evaluated whether interactions between the PGS_LDL_ and change in saturated fat intake differed by fatty acid chain length. Of the 11 saturated fatty acids available, we examined the three that each contributed at least 1% of total energy at baseline: myristic acid (C14:0), palmitic acid (C16:0), and stearic acid (C18:0), which together made up 88% of total saturated fat intake (at baseline and 6 months). Baseline and 6 month mean intakes are shown in **Supplementary Figure 1A**. All three fatty acids showed significant PGS_LDL_-by-change interactions after Bonferroni correction with similar directions and patterns of associations across tertiles for all three fatty acids (all *P* interaction < 0.01; **Supplementary Figure 1B**), consistent to those observed in the total saturated fat analysis (**Figure 3A**).

### Interaction between the polygenic score and changes in other macronutrients

We further evaluated interactions between the PGS_LDL_ and changes in other macronutrients, including unsaturated fat, protein and carbohydrate, on changes in LDL-C. Of these, none showed a significant interaction in either diet group (**Supplementary Figure 2**). These findings suggest that the observed gene-diet interaction was specific to changes in saturated fat intake in the HLC group.

## Discussion

In this secondary analysis of the DIETFITS trial, we found that individuals with a genetic propensity towards high LDL-C were at increased risk for LDL-C elevation and new hypercholesterolemia after 6 months on a HLC diet, independent of baseline LDL-C, weight change, and other factors. Further, some of this risk may be attributed to an interaction between saturated fat intake and polygenic background. In contrast, the LDL-C lowering effects of the HLF diet were less impacted by genetics. Together, these findings suggest that individuals with a genetic propensity towards high LDL-C may be more sensitive to the adverse effects of saturated fat in the context of low-carbohydrate diets.

Our findings extend existing evidence in several important ways. In the UK Biobank, a self-reported low-carbohydrate, high fat dietary pattern (<25% of total energy from carbohydrate and > 45% from fat) was associated with higher LDL-C only among participants in the 20% of polygenic risk for hypercholesterolemia (16). Within this group, severe hypercholesterolemia (LDL-C > 190 mg/dL) was three times more prevalent among those reporting the low- carbohydrate, high fat diet compared with those reporting a standard diet (16). In the present study, we extend these findings in a randomized dietary intervention that achieved a comparable macronutrient distribution (26% carbohydrate, 47% fat). We found that each standard deviation increase in the polygenic score was associated with more than twice the odds of developing hypercholesterolemia after 6 months on a HLC diet, independent of baseline LDL-C and weight change. Importantly, we further demonstrate that this genetic susceptibility was partly attributable to greater sensitivity to increases in saturated fat intake. These findings are consistent with our recent observational analysis in the Women’s Health Initiative, in which we found that the association of statured fat intake and LDL-C was amplified among individuals with higher polygenic risk for elevated LDL-C, with similar patterns extending to future risk of atherosclerotic cardiovascular disease (15). Collectively, findings across distinct populations and complementary study designs strengthen the evidence that polygenic background contributes meaningfully to interindividual differences in the LDL-C response to low-carbohydrate diets, in part through genetically driven differences in sensitivity to saturated fat.

The mechanism underlying the presence of genetic effects on the HLC diet is uncertain, but the interaction with saturated fat, together with the absence of interactions with other macronutrients, points to a plausible explanation. Mechanistic studies in animal models have shown that saturated fat intake raises LDL-C by suppressing hepatic LDL receptor activity, thereby reducing receptor-mediated LDL clearance (32–34). Human tracer studies have similarly reported slower LDL clearance with saturated fat compared to polyunsaturated fat (35), and controlled feeding studies show that reducing saturated fat or replacing it with unsaturated fat upregulates LDL receptor expression in circulating mononuclear cells (14, 36, 37). Polygenic scores for LDL-C capture, in part, common variation at loci governing LDL clearance. Individuals with a higher genetic burden may therefore be less able to compensate for diet-induced reductions in LDL clearance, resulting in a larger increase for a given rise in saturated fat intake. Future studies are needed to understand the pathways through which the PGS_LDL_ operates.

Saturated fatty acids vary in carbon chain length and have been reported to have distinct effects on LDL-C. Dietary intervention trials have shown that palmitic and myristic acids generally raise LDL-C when substituted for carbohydrate or unsaturated fat, whereas stearic appears to have little effect (38, 39). In our cohort, palmitic, myristic and stearic made up 88% of saturated fat intake at baseline and 6 months, with palmitic contributing to more than half. In the HLC group, changes in all three fatty acids were associated with increases in LDL-C in the top two tertiles of polygenic risk, but not among those in the lowest tertile. The association of stearic acid was unexpected based on prior evidence (38, 39), and may reflect the substantial overlap in food sources of these fatty acids, particularly meat and dairy, making their individual effects difficult to distinguish in this setting. Further investigation in controlled feeding trials that can better isolate the effects of the individual fatty acids is warranted. Alternatively, examining specific food sources of saturated fat may provide additional insight, as the food matrix in which the fatty acids are consumed may also contribute to their differential effects on LDL-C (40).

In contrast to the HLC arm, the polygenic score did not predict LDL-C response to the HLF diet. This finding is consistent with a prior analysis of a subset of 145 women in the intervention arm of the Dietary Modification Trial of the Women’s Health Initiative, in which a main-effect polygenic score for LDL-C did not predict 1-year LDL-C response to a low-fat dietary intervention, whereas a diet-response polygenic score did, explaining 3.7% of the variance in LDL-C change (41). Together, these results suggest that the genetic variants that predispose individuals to higher LDL-C concentrations may not translate proportionally into responsiveness to fat reduction. This responsiveness may be better captured by scores specifically derived from gene-diet interactions. Dietary context may also play a role. Both low-fat dietary interventions were multifactorial and emphasized fiber-rich foods, including fruits, vegetables and whole grains. This dietary pattern is known to support favorable LDL-C levels (30) and may have attenuated genetically mediated responses. Additional research is needed to determine whether overall dietary context shapes the expression of genetic effects on LDL-C response, and whether diet-response polygenic scores offer greater predictive utility across different dietary interventions.

Our findings have important clinical implications. Substantial interindividual variability in LDL-C response to low-carbohydrate diets has been widely documented (4), and our results suggest that this variability is not random but is explained, in part, by polygenic risk for elevated LDL-C. Because conflicting nutrition messages can undermine dietary advice (42, 43), acknowledging that LDL-C responses differ for biologically meaningful reasons may help strengthen clinician-patient trust. Our findings also counter the notion that diet matters little for individuals genetically predisposed to elevated LDL-C. Rather, these individuals appear to be most susceptible to LDL-C increases on a low-carbohydrate diet, in part through heightened sensitivity to saturated fat intake. Because these diets are often adopted for weight loss or glycemic control, rather than lipid management, such LDL-C increases may go unrecognized. Although polygenic scores are not yet ready for clinical use, our findings reinforce recommendations to measure LDL-C before and after initiating low-carbohydrate, high-fat diets (4). For those whose preferences and lifestyles align with this dietary pattern, replacing saturated fat with unsaturated fat remains a guideline-recommended strategy (30, 31).

Our findings should be interpreted in light of several limitations. First, this study was a post hoc analysis of a secondary endpoint of the DIETFITS trial. Second, diet was self-reported, and variable adherence may have contributed to heterogeneity in response. However, adherence at 6 months was generally good, as supported by repeated, unannounced 24-hour recalls (17), and findings remained consistent after adjusting for adherence. Third, although diet assignment was randomized, changes in saturated fat intake within each arm were not. Nonetheless, evaluating within-person changes in the context of randomly assigned interventions reduced the potential for confounding by diet self-selection, time-invariant factors, and reverse causation. Fourth, the study population consisted of individuals with overweight or obesity, which may limit generalizability. This is particularly relevant given evidence that the effect of saturated fat on LDL-C differs between individuals with and without obesity (44). Fifth, both diet interventions emphasized high diet quality, allowing us to evaluate these associations within the context of relatively healthful dietary intake. Whether these findings extend to lower-quality or more extreme versions of these dietary patterns remains unclear. Sixth, most participants were of European ancestry, and the limited sample sizes in other genetic ancestry groups precluded ancestry-stratified analyses, limiting out ability to assess whether findings were consistent across all ancestry groups.

In conclusion, genetic effects help explain the heterogeneity in LDL-C response to a HLC diet. Individuals with a high polygenic score for LDL-C have increased risk for an adverse LDL-C response to this dietary pattern. This risk may be partly explained by heightened sensitivity to saturated fat intake in the context of a low-carbohydrate diet.

## Supporting information

Supplemental Content

## Author contributions

AB and SLC designed the present study and drafted the manuscript. AB conducted the analyses. CDG designed and led the DIETFITS trial, collected the data, and edited the manuscript. All authors reviewed and approved the final manuscript.

## Data Sharing Statement

Data pertaining to this analysis will be shared for approved study proposals. Data use agreement is required.

## Declaration of Generative AI and AI-assisted technologies in the writing process

The authors declare that no generative AI or AI-assisted technologies were used in the writing of this manuscript.

