## Supplemental Content for "Genetic Risk and LDL-C Response to Low-Carbohydrate and Low-Fat Diets: A Secondary Analysis of the DIETFITS Trial"

**Supplementary Table 1. Dietary intake by time point.[1]**

| | Healthy Low-Carbohydrate Diet (n = 223) | Healthy Low-Fat Diet (n = 208) |
|---|---|---|
| **Total energy, kcal** | | |
| Baseline | 2214 (626.9) | 2164 (674.1) |
| 6 months | 1644.5 (528.3) | 1653.8 (590.9) |
| Change | -569.5 (649.1) | -510.0 (654.9) |
| **Carbohydrates, % kcal** | | |
| Baseline | 44.2 (7.2) | 44.4 (7.6) |
| 6 months | 26.1 (10.5) | 50.99 (10.5) |
| Change | -18.1 (11.6) | 6.59 (10.5) |
| **Total fat, % kcal** | | |
| Baseline | 35.9 (5.8) | 35.2 (6.1) |
| 6 months | 47.4 (8.7) | 26.5 (8.7) |
| Change | 11.5 (9.6) | -8.7 (9.7) |
| **Saturated fat, % kcal** | | |
| Baseline | 11.9 (2.9) | 11.6 (2.9) |
| 6 months | 15.1 (4.0) | 8.0 (3.5) |
| Change | 3.2 (4.5) | -3.6 (3.9) |
| **Unsaturated fat, % kcal** | | |
| Baseline | 21.0 (3.9) | 20.7 (4.3) |
| 6 months | 28.1 (6.4) | 16.1 (5.6) |
| Change | 7.1 (7.2) | -4.6 (6.6) |
| **Protein, % kcal** | | |
| Baseline | 17.3 (3.7) | 17.7 (4.1) |
| 6 months | 24.1 (6.3) | 20.5 (5.6) |
| Change | 6.9 (6.3) | 2.8 (5.7) |
| **Alcohol, % kcal** | | |
| Baseline | 2.9 (4.4) | 2.9 (4.5) |
| 6 months | 2.5 (4.5) | 2.3 (3.9) |
| Change | -0.4 (5.4) | -0.6 (4.6) |
| **Fiber, grams** | | |
| Baseline | 21.8 (8.5) | 22.4 (9.6) |
| 6 months | 17.6 (7.2) | 24.1 (10.4) |
| Change | -4.1 (8.9) | 1.6 (10.9) |
| **Healthy Eating Index-2010** | | |
| Baseline | 55.1 (12.0) | 56.7 (11.0) |
| 6 months | 59.9 (11.0) | 64.4 (12.7) |
| Change | 4.7 (13.7) | 7.7 (12.2) |

[1]Values are presented as the mean (SD). Healthy Eating Index-2010 values range from 0 to 100, with higher scores indicating better diet quality.

**Supplementary Table 2. Association between the polygenic score for LDL-C and baseline LDL-C levels by diet group.[1]**

| | **Healthy Low-Carbohydrate Diet (n = 223)** | **Healthy Low-Fat Diet (n = 208)** |
|---|---|---|
| β (SE) | 10.3 (1.6) | 12.7 (2.0) |
| *P* value | < 0.0001 | < 0.0001 |
| Incremental $R^2$ (95% CI), % | 14.7 (5.4, 21.9) | 17.0 (6.8, 28.4) |

Abbreviations: LDL-C, low-density lipoprotein cholesterol; $PGS_{LDL}$, polygenic score for low-density lipoprotein cholesterol.
[1]The β coefficients (SE) are from multivariable linear regression models with baseline LDL-C as the response variable and $PGS_{LDL}$ as the predictor variable. Models are adjusted for age, sex, and genetic principal components 1-3. Incremental $R^2$ represents additional percentage of variance in baseline LDL-C explained by $PGS_{LDL}$ beyond the base covariate model, calculated as the difference in $R^2$ between models with and without $PGS_{LDL}$. The 95% CIs were estimated using bootstrapping.

**Supplementary Table 3. Distribution of genetically inferred ancestry groups across polygenic score tertiles in the full study cohort and by diet group.**[1]

| All Participants (n = 431) | | | | | |
|---|---|---|---|---|---|
| | | $PGS_{LDL}$ Tertile | | | |
| **Genetically Inferred Ancestry** | **n** | **Low (%)** | **Middle (%)** | **High (%)** | ***P* value** |
| European | 272 | 90 (62.1) | 94 (65.7) | 88 (61.5) | 0.75 |
| American | 87 | 29 (20.0) | 30 (21.0) | 28 (19.6) | |
| East Asian | 31 | 11 (7.6) | 6 (4.2) | 14 (9.8) | |
| Central/South Asian | 27 | 10 (6.9) | 7 (4.9) | 10 (7.0) | |
| African | 14 | 5 (3.4) | 6 (4.2) | 3 (2.1) | |
| **Healthy low-carbohydrate diet (n = 223)** | | | | | |
| | | $PGS_{LDL}$ Tertile | | | |
| **Genetically Inferred Ancestry** | **n** | **Low (%)** | **Middle (%)** | **High (%)** | ***P* value** |
| European | 145 | 44 (58.7) | 50 (67.6) | 51 (68.9) | 0.83 |
| American | 39 | 14 (18.7) | 14 (18.9) | 11 (14.9) | |
| East Asian | 19 | 8 (10.7) | 4 (5.4) | 7 (9.5) | |
| Central/South Asian | 14 | 7 (9.3) | 4 (5.4) | 3 (4.1) | |
| African | 6 | 2 (2.7) | 2 (2.7) | 2 (2.7) | |
| **Healthy low-fat diet (n = 208)** | | | | | |
| | | $PGS_{LDL}$ Tertile | | | |
| **Genetically Inferred Ancestry** | **n** | **Low (%)** | **Middle (%)** | **High (%)** | ***P* value** |
| European | 127 | 46 (65.7) | 44 (63.8) | 37 (53.6) | 0.35 |
| American | 48 | 15 (21.4) | 16 (23.2) | 17 (24.6) | |
| East Asian | 12 | 3 (4.3) | 2 (2.9) | 7 (10.1) | |
| Central/South Asian | 13 | 3 (4.3) | 3 (4.3) | 7 (10.1) | |
| African | 8 | 3 (4.3) | 4 (5.8) | 1 (1.4) | |

Abbreviations: $PGS_{LDL}$, polygenic score for low-density lipoprotein cholesterol.
[1]Values are reported as n (%). *P* values from a chi-squared test comparing genetically inferred ancestry across $PGS_{LDL}$ tertiles.

**Supplementary Table 4. Associations between polygenic score and 6-month LDL-C response in the healthy low-carbohydrate diet group.**[1]

| | | LDL-C increase[2] | | | New hypercholesterolemia[3] | | |
|---|---|---|---|---|---|---|---|
| Subset | Model | n | OR [95% CI] | *P* | n | OR [95% CI] | *P* |
| All | Model 1 | (↑) 127 (↓) 96 | 1.43 [1.02, 2.05] | 0.041 | (yes) 23 (no) 128 | 2.27 [1.18, 4.81] | 0.021 |
| | Model 2 | (↑) 127 (↓) 96 | 1.44 [1.03, 2.06] | 0.039 | (yes) 23 (no) 128 | 2.28 [1.18, 4.84] | 0.021 |
| | Model 3 | (↑) 127 (↓) 96 | 1.44 [1.03, 2.06] | 0.039 | (yes) 23 (no) 128 | 2.28 [1.18, 4.87] | 0.020 |
| Subset | Model | n | OR [95% CI] | *P* | n | OR [95% CI] | *P* |
| EUR only | Model 1 | (↑) 86 (↓) 59 | 1.94 [1.22, 3.24] | 0.008 | (yes) 14 (no) 83 | 2.98 [1.20, 10.22] | 0.039 |
| | Model 3 | (↑) 86 (↓) 59 | 1.96 [1.22, 3.27] | 0.007 | (yes) 14 (no) 83 | 2.97 [1.19, 10.27] | 0.041 |
| | Model 3 | (↑) 86 (↓) 59 | 1.96 [1.22, 3.27] | 0.007 | (yes) 14 (no) 83 | 3.68 [1.31, 16.76] | 0.036 |

Abbreviations: EUR, European ancestry; LDL-C, low-density lipoprotein cholesterol; $PGS_{LDL}$, polygenic score for low-density lipoprotein cholesterol.

[1]Odds ratios (OR) and 95% confidence intervals (CI) are from logistic regression models and represent the change in odds per 1-SD increase in the $PGS_{LDL}$. Model 1 was adjusted for age, sex, baseline LDL-C , 6-month weight change, 6-month change in saturated fat intake, and genetic principal components 1-3. Model 2 was adjusted for Model 1 covariates plus a modified Healthy Eating Index-2010 that excluded the fatty acid and solid fats components. Model 3 was adjusted for Model 2 covariates plus dietary adherence.

[2]For the LDL-C direction of response analysis, the outcome was an increase versus decrease in LDL-C from baseline to 6 months; total sample sizes were 223 participants overall and 145 participants of European ancestry.

[3]For the new hypercholesterolemia analysis, the outcome was new-onset hypercholesterolemia defined as LDL-C ≥130 mg/dL at 6 months among participants with normal baseline LDL-C (LDL-C < 130 mg/dL); total sample sizes were 151 participants overall and 97 participants of European ancestry.

**Supplementary Table 5. Associations between polygenic score and 6-month LDL-C response in the healthy low-fat diet group.**[1]

| | | LDL-C decrease | | |
|---|---|---|---|---|
| Subset | Model | n | OR [95% CI] | *P* |
| All | Model 1 | (↓) 130 (↑) 78 | 0.85 [0.59, 1.21] | 0.38 |
| | Model 2 | (↓) 130 (↑) 78 | 0.85 [0.59, 1.21] | 0.37 |
| | Model 3 | (↓) 130 (↑) 78 | 0.85 [0.59, 1.21] | 0.38 |
| Subset | Model | n | OR [95% CI] | *P* |
| EUR only | Model 1 | (↓) 88 (↑) 39 | 0.90 [0.53, 1.48] | 0.69 |
| | Model 3 | (↓) 88 (↑) 39 | 0.88 [0.51, 1.47] | 0.63 |
| | Model 3 | (↓) 88 (↑) 39 | 0.91 [0.52, 1.53] | 0.72 |

Abbreviations: EUR, European ancestry; LDL-C, low-density lipoprotein cholesterol; $PGS_{LDL}$, polygenic score for low-density lipoprotein cholesterol.
[1]Odds ratios (OR) and 95% confidence intervals (CI) are from logistic regression models and represent the change in odds per 1-SD increase in the $PGS_{LDL}$. Model 1 was adjusted for age, sex, baseline LDL-C , 6-month weight change, 6-month change in saturated fat intake, and genetic principal components 1-3. Model 2 was adjusted for Model 1 covariates plus a modified Healthy Eating Index-2010 that excluded the fatty acid and solid fats components. Model 3 was adjusted for Model 2 covariates plus dietary adherence. The outcome was a decrease vs increase in LDL-C from baseline to 6 months; total sample sizes were 208 participants overall and 127 participants of European ancestry.

**Supplementary Table 6. Sensitivity analyses examining the interaction between polygenic score for LDL-C and change in saturated fat intake on 6-month change in LDL-C.[1]**

| | | Healthy Low-Carbohydrate Diet (n = 223) | | Healthy Low-Fat Diet (n = 208) | |
|---|---|---|---|---|---|
| **Model** | **Independent variable** | **β (SE)** | **P value** | **β (SE)** | **P value** |
| Model 1 | Δ SFA (1 % kcal) | 1.03 (0.28) | 0.0003 | 1.23 (0.34) | 0.0003 |
| | $PGS_{LDL}$ (1-SD) | 3.17 (1.65) | 0.056 | 5.93 (1.98) | 0.003 |
| | $PGS_{LDL}$ x Δ SFA | 0.69 (0.23) | 0.003 | 0.56 (0.36) | 0.12 |
| Model 2 | Δ SFA (1 % kcal) | 1.03 (0.28) | 0.0003 | 1.24 (0.34) | 0.0003 |
| | $PGS_{LDL}$ (1-SD) | 3.17 (1.66) | 0.057 | 5.90 (1.99) | 0.003 |
| | $PGS_{LDL}$ x Δ SFA | 0.69 (0.23) | 0.003 | 0.56 (0.36) | 0.12 |
| Model 3 | Δ SFA (1 % kcal) | 1.08 (0.29) | 0.0003 | 1.30 (0.36) | 0.0004 |
| | $PGS_{LDL}$ (1-SD) | 3.23 (1.66) | 0.053 | 5.93 (1.99) | 0.003 |
| | $PGS_{LDL}$ x Δ SFA | 0.68 (0.23) | 0.003 | 0.57 (0.36) | 0.12 |

Abbreviations: LDL-C, low density lipoprotein cholesterol; $PGS_{LDL}$, polygenic score for LDL-C, Δ SFA, change in saturated fat intake (as percent of total daily energy) from baseline to 6 months.
[1] The β coefficients (SE) represent the change in LDL-C associated with a 1-SD increase in the $PGS_{LDL}$ or a 1-percentage-point increase in SFA from baseline to 6 months. Within each model, the main effects of $PGS_{LDL}$ and Δ SFA and their interaction term were estimated simultaneously in the same multivariable regression model. Model 1 was adjusted for age, sex, baseline LDL-C , 6-month weight change, and genetic principal components 1-3. Model 2 was adjusted for Model 1 covariates plus a modified Healthy Eating Index-2010 that excluded the fatty acid and solid fats components. Model 3 was adjusted for Model 2 covariates plus dietary adherence.

**Supplementary Table 7. Sensitivity analyses for the interaction models of polygenic score and change in saturated fat intake on 6-month change in LDL-C among individuals of genetically inferred European ancestry.**[1]

| | | Healthy Low-Carbohydrate Diet (n = 145) | | Healthy Low-Fat Diet (n = 127) | |
|---|---|---|---|---|---|
| **Model** | **Independent variable** | **β (SE)** | **P value** | **β (SE)** | **P value** |
| Model 1 | Δ SFA (1 % kcal) | 1.08 (0.33) | 0.0015 | 1.00 (0.40) | 0.013 |
| | $PGS_{LDL}$ (1-SD) | 3.49 (2.16) | 0.11 | 4.87 (2.60) | 0.063 |
| | $PGS_{LDL}$ x Δ SFA | 0.64 (0.25) | 0.012 | 0.38 (0.42) | 0.36 |
| Model 2 | Δ SFA (1 % kcal) | 1.08 (0.34) | 0.0015 | 1.00 (0.40) | 0.013 |
| | $PGS_{LDL}$ (1-SD) | 3.42 (2.17) | 0.12 | 4.86 (2.62) | 0.066 |
| | $PGS_{LDL}$ x Δ SFA | 0.64 (0.25) | 0.013 | 0.38 (0.42) | 0.36 |
| Model 3 | Δ SFA (1 % kcal) | 1.20 (0.34) | 0.0006 | 0.78 (0.44) | 0.081 |
| | $PGS_{LDL}$ (1-SD) | 3.27 (2.16) | 0.13 | 4.59 (2.62) | 0.083 |
| | $PGS_{LDL}$ x Δ SFA | 0.65 (0.25) | 0.012 | 0.35 (0.42) | 0.40 |

Abbreviations: LDL-C, low density lipoprotein cholesterol; $PGS_{LDL}$, polygenic score for LDL-C, Δ SFA, change in saturated fat intake (as percent of total daily energy) from baseline to 6 months.
[1]The β coefficients (SE) represent the change in LDL-C associated with a 1-SD increase in the $PGS_{LDL}$ or a 1-percentage-point increase in SFA from baseline to 6 months. Within each model, the main effects of $PGS_{LDL}$ and Δ SFA and their interaction term were estimated simultaneously in the same multivariable regression model. Model 1 was adjusted for age, sex, baseline LDL-C , 6-month weight change, and genetic principal components 1-3. Model 2 was adjusted for Model 1 covariates plus a modified Healthy Eating Index-2010 that excluded the fatty acid and solid fats components. Model 3 was adjusted for Model 2 covariates plus dietary adherence.

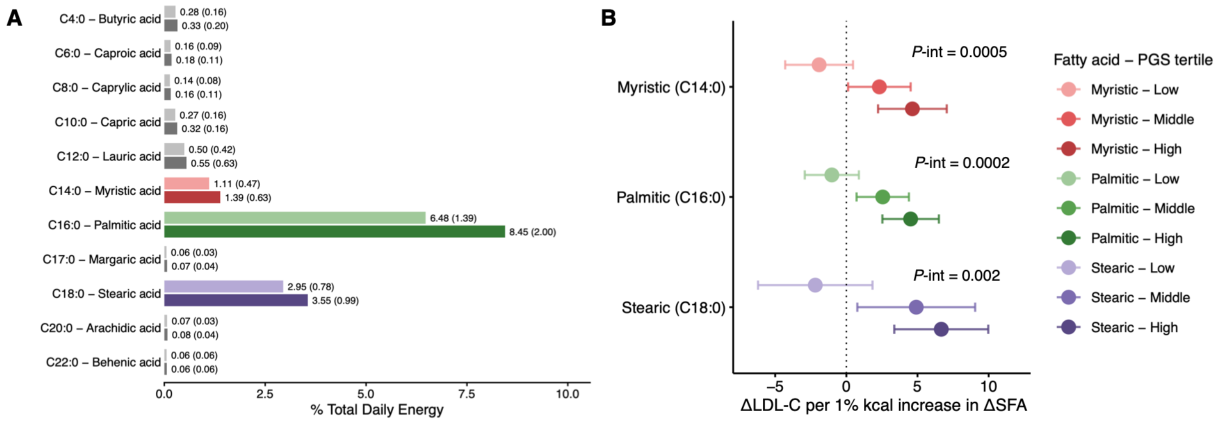


**Supplementary Figure 1. Association of change in saturated fatty acid intake with change in LDL-C according to polygenic score tertile in the healthy low-carbohydrate diet group.** (A) Mean (SD) percentage of total daily energy intake from individual saturated fatty acids at baseline (light bars) and 6 months (dark bars). (B) Estimated change in low-density lipoprotein cholesterol (LDL-C) per 1-percentage-point increase in saturated fatty acid intake (SFA) from baseline to 6 months stratified by $PGS_{LDL}$ tertiles. Points represent β coefficients and error bars denote 95% CIs from multivariable linear regression models adjusted for age, baseline LDL-C, 6-month weight change, and genetic principal components 1-3. *P* values (*P*-int) indicate interaction between change in saturated fatty acid intake and $PGS_{LDL}$ tertile. $PGS_{LDL}$ , polygenic score for LDL-C.

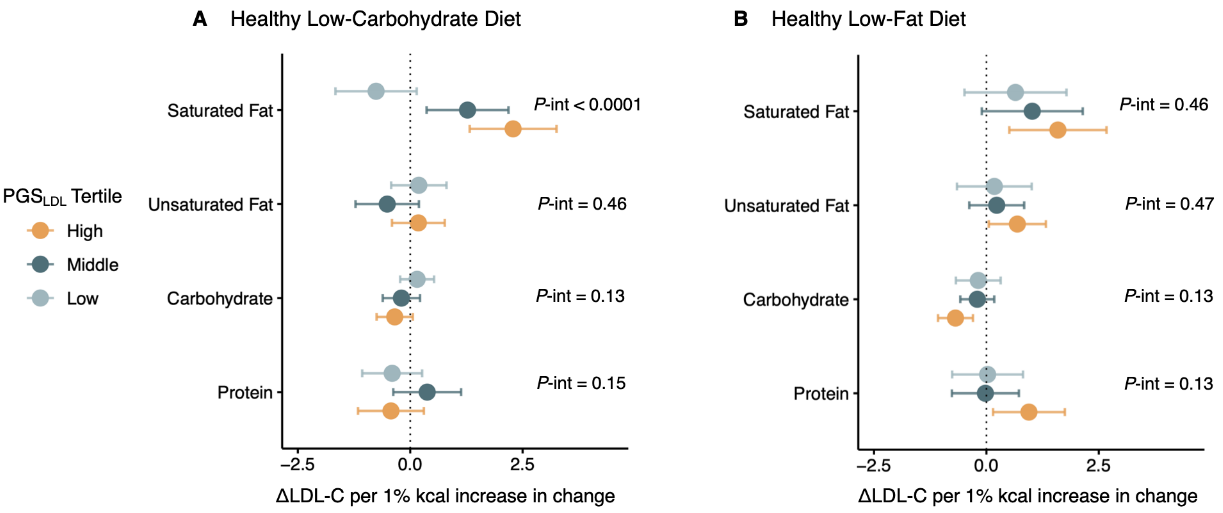


**Supplementary Figure 2. Associations of changes in macronutrient intake with change in LDL-C according to polygenic score tertile by diet group.** Estimated change in low-density lipoprotein cholesterol (LDL-C) per 1-percentage-point increase in each macronutrient from baseline to 6 months stratified by $PGS_{LDL}$ tertiles, among participants randomized to the healthy-low-carbohydrate diet (A) and healthy low-fat diet (B). Points represent β coefficients and error bars denote 95% CIs from multivariable linear regression models adjusted for age, baseline LDL-C, 6-month weight change, and genetic principal components 1-3. Effect estimates are presented as per 1-percentage-point increase in each macronutrient, irrespective of the average direction of change within each diet group; mean changes in macronutrient intake by diet group are shown in Supplementary Table 1. *P* values (*P*-int) indicate interaction between change in macronutrient intake and $PGS_{LDL}$ tertile. $PGS_{LDL}$ , polygenic score for LDL-C.
